# Spring Weather and COVID-19 Deaths in the U.S.

**DOI:** 10.1101/2020.06.20.20136259

**Authors:** Seyed M. Karimi, Mahdi Majbouri, Kelsey White, Bert Little, W. Paul McKinney, Natalie DuPre

## Abstract

This study used statistically robust regression models to control for a large set of confounders (including county-level time-invariant factors and time trends, regional-level daily variation, state-level social distancing measures, ultraviolet light, and levels of ozone and fine particulate matter, PM2.5) to estimate a reliable rather than simple regression for the impact of weather on the most accurately measured outcome of COVID-19, death. When the average minimum temperature within a five-day window increased by one degree Fahrenheit in spring 2020, daily death rates in northern U.S. counties increased by an estimated 5.1%. When ozone concentration over a five-day window rose by one part per billion, daily death rates in southern U.S. counties declined by approximately 2.0%. Maximum temperature, precipitation, PM2.5, and ultraviolet light did not significantly associate with COVID-19 mortality. The mechanism that may drive the observed association of minimum temperature on COVID-19 deaths in spring months may be increased mobility and contacts. The effect of ozone may be related to its disinfectant properties, but this requires further confirmation.

## Introduction

The effect of weather changes is important to understand, especially changes in temperature, on COVID-19 infections, hospitalizations, and deaths. Knowledge of the effect of weather conditions on SARS-CoV-2 (the virus causing Covid-19) can inform health care systems’ preparations at local, state, and national levels. In addition, by studying the relationship between environmental factors and COVID-19 mortality, we may learn more about how environmental factors influence the spread and transmission of the disease to generate novel hypotheses on transmission patterns and preventive strategies.

Associations between weather and viral prevalence, population incidence rates, or virus survival time were explored prior to the appearance of COVID-19 in human populations. Temperature and humidity modulate the transmission and survival of influenza viruses.^1–3^ The transmission of the H1N1 flu virus in 2009 increased in areas with lower absolute humidity and open schools.^4^ Viruses related to SARS-CoV-2 (e.g., SARS-CoV-1 between 2002 and 2003) increased in prevalence for areas with lower temperatures, a wider range between maximum and minimum temperatures, and higher wind velocity.^5^ The highest prevalence occurred when temperatures averaged 62.4°Fahrenheit (95% Confidence Interval, CI: 51.3°F−88.0°F), and relative humidity averaged 52.2% (95% CI, 33.0%−71.4%).^5^ Also, higher temperatures and higher levels of ultraviolet (UV) light were associated with higher MERS-CoV transmission rates, a virus similar to SARS-CoV-2.^6,7^

Transmission of a virus from one person to another requires social proximity, favorable environmental conditions, and host susceptibility. Increased physical interactions and social behavior influence the transmission of various viral infections.^8^ Influenza transmission is higher on workdays compared to weekends and holidays, apparently due to increased physical and social contacts and the duration of contact.^3,9^ An early study of SARS-CoV-2 transmission in Wuhan highlighted the possibility of higher transmissibility due to an increased population density.^10^ In addition, environmental factors impact viral transmission. Ambient pollutants negatively impacting the immune system and may make individuals who are exposed to pollutants more susceptible to infectious diseases.^11^ Areas with higher concentrations of particulate matter appear to sustain SARS-CoV-2 for longer,^12^ and long-term exposure could increase mortality rates.^13^ Also, weather conditions affect how long a virus survives on surfaces and in the air. The survival time of a virus on a surface depends on its exposure to temperature, humidity, and concentration.^14^ As the temperature approaches 86°F and relative humidity decreases, the shorter the survival time of SARS-CoV-2 and similar viruses.^14– 17^ The survival time of SARS-CoV-2 in a person’s nasal sputum and mucosa decreases as the relative humidity and temperature increase. The virus can remain viable for at least 7 days, even in warm, humid conditions.^17^ The stability of SARS-CoV-2 in aerosols and conditions similar to the upper respiratory tract led to a warning that its transmission may exponentially increase.^18^ Host susceptibility depends, on the other hand, on individual health status, genetic, and other contextual factors. Geographical regions that expose populations to various antigens and higher levels of vitamin D through sunlight may increase a population’s inherent immunity to the novel coronavirus.^19^

Analyses of how weather influences SARS-CoV-2 transmission have yielded mixed results. Some findings suggest that the virus may be similar to a seasonal respiratory virus where the higher the temperature, the lower the transmission rate.^10,20–26^ Transmissions rates also appear lower when humidity is higher^20–31^ and when an area has a higher UV light index.^23,26^ Reductions in transmission also appear as wind speed and precipitation decrease.^20,25,32^ Other studies, however, reported weak or inconclusive relationships with weather, ^23,33–35^ which warranted a cautious interpretation.^36,37^

Inconclusive research on the effect of weather and environmental factors on SARS-CoV-2 transmission may emerge from methodological challenges such as utilizing only a limited number of environmental factors^21,22,30,31,34,38,39^ without controlling for important influences such as government mitigation efforts, public responses, population density, and local practices.^20,22,23,26,27,30,40,41^ Also, COVID-19 research that examined COVID-19 cases suffers from potentially substantial measurement error from lack of systematic representative testing, leading to uncertainty about the true prevalence and incidence COVID-19 in study communities. While the total case estimates are underestimates of the truth, measurement error can be lowered by using hard outcomes such as the number of COVID-19 deaths, especially if deaths occurred in a hospital, frequently the case in the United States. Research on environmental and meteorological effects on COVID-19 fatality is scarce. Two studies mapped global COVID-19 deaths against the global pattern of climate changes and predicted that COVID-19 deaths increase as the weather becomes warmer in spring.^42,43^ Two time-series studies explored the relationship between weather changes and COVID-19 deaths for specific cities in China and found positive predictive associations.^44,45^ On the other hand, among the two studies that examined cross-national variation in COVID-19 deaths by temperature and precipitation, one found no association^46^ and another found a negative association with COVID-19 death.^47^ Lastly, one study that used the average long-term levels of weather and air pollutants across U.S. counties found no association between COVID-19 deaths and a county’s average summer and winter temperatures, but found a positive association between that and the historical PM_2.5_ levels.^48^

It is important for literature on environmental factors and COVID-19 mortality to carefully consider the influence of confounding factors. First, using country fixed-effects as well as time fixed-effects are necessary to capture many potential correlates. Some studies controlled for country fixed-effects but only included a weak form of time fixed-effects or sometimes, only time trends.^26,46,47^ Thus, non-linear trends and measurable and unmeasurable changes in government policies were not captured. Additionally, much of the literature did not control for serial correlation in cases, deaths, and weather variables.^26,46,47^ Serial correlation can result in underestimates of coefficient standard errors hence produces spuriously statistically significant results. Furthermore, data quality across countries and over time varies due to variation in testing capacity, lack of resources or knowledge to identify COVID deaths correctly. Assessing non-linear trends over time due to testing capacity and health care resources and knowledge that differ across geography, and hence environmental factors, is important to consider when studying environmental factors and COVID-19 outcomes.

This study used county-level COVID-19 death numbers across the United States and examined their association with minimum and maximum daily temperatures at the approximate time of exposure to SARS-CoV-2. To address the aforementioned issues of confounding, serial correlation, and time trends, this analysis accounted for time-constant factors, linear and non-linear time-varying factors, serial correlation, social distancing measures, and daily levels of precipitation, ozone, PM2.5, and UV light.

## Methods

### Data Assembling Procedure

Data from seven different sources were integrated to assemble the work file used in this analysis. Daily county-level COVID-19 cases and deaths data, collected from state and local public health departments and compiled by the New York Times from the beginning of the pandemic in the U.S. until May 16, 2020, were extracted.^49^ New York Times complies the data from state and local public health departments on a daily basis. County-level COVID-19 data were geocoded using the U.S. county centroid latitude and longitude provided by the National Weather Service of the National Oceanic and Atmospheric Administration (NOAA).^50^ Next, the geocoded COVID-19 dataset was supplemented with county population characteristics (i.e., the population by age group, gender, race, and ethnicity) from the U.S. Census Bureau.^51^ The Bureau’s latest (2018) estimations of the U.S. counties’ population characteristics were used. Four state-level social distancing measures were added to the dataset that contained facilities closures (i.e., when a shelter-in-place order was issued, gatherings of 500 or more were banned, public schools were closed, restaurants, gyms, entertainment facilities) (**Figure 1**).^52^

**Figure 1.**
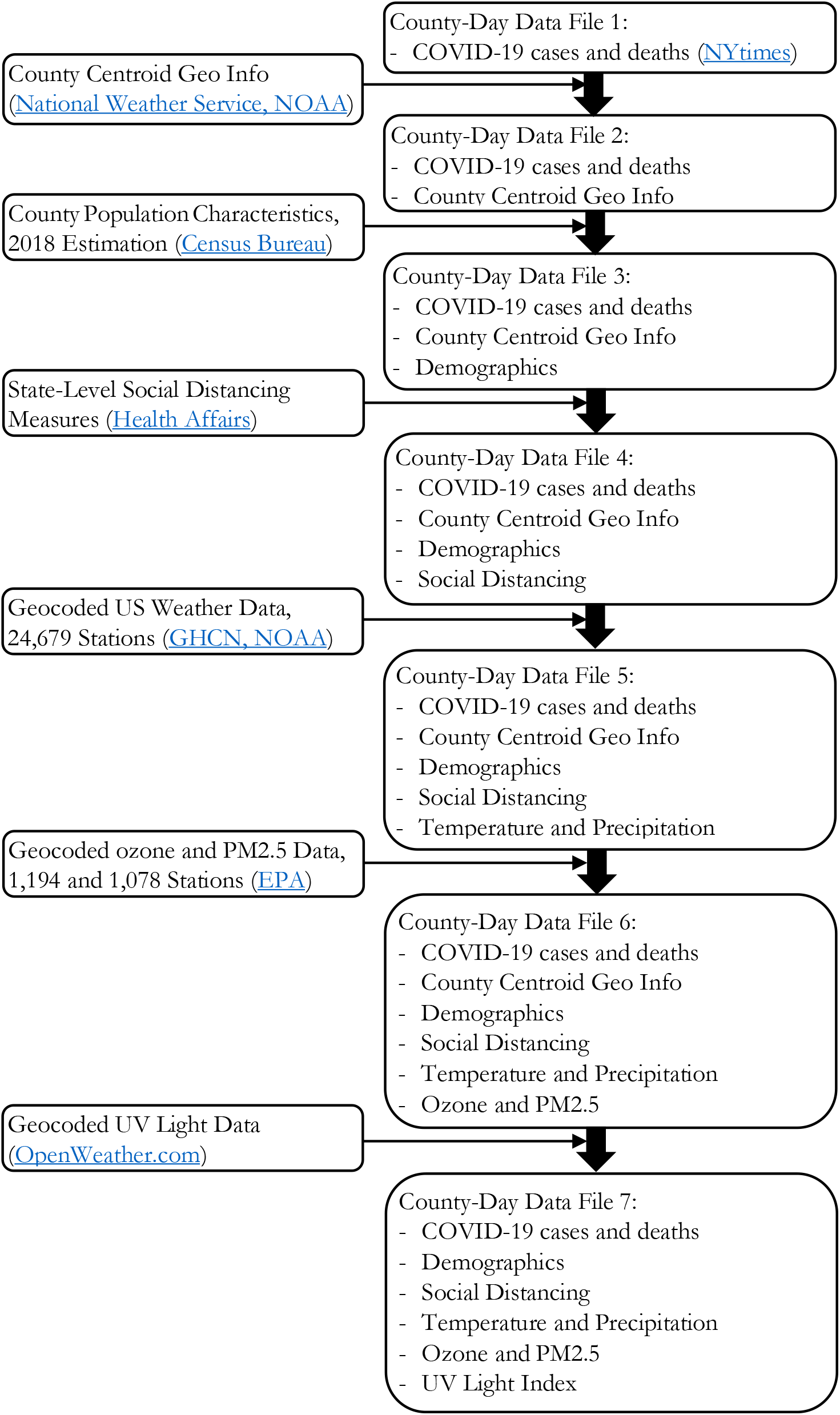
Data assembling procedure

NOAA’s Global Historical Climatology Network (GHCN)-Daily data were added to supplement COVID-19 and population data,^53^ and included temperature (daily maximum and minimum), precipitation, snowfall, and snow depth-labeled as core meteorological elements. Additional meteorological information (e.g., average daily temperature, evaporation, wind speed, wind direction, and cloudiness) was not used because it included many missing values that required estimation by interpolation. Daily data on the core meteorological elements from 24,679 US weather stations from January 1 to May 16, 2020, were combined. Each county-day with the core weather information recorded by the first to the tenth nearest station to the county’s centroid. More than one station was used for a county to facilitate estimation of county’s nearest station to estimate missing values with a linear estimate of surrounding stations.

Air pollution information was obtained from the U.S. Environmental Protection Agency (EPA)’s outdoor air quality data portal,^54^ which is pollutant-specific. Ozone (O3) and PM2.5 (fine particulate matter) from 2020 that had substantially less missing values than other pollutants gathered by the EPA for 2020 were selected. The influence of the two air pollutants on SARS-CoV-2 has been studied.^12,48,55,56^ Daily maximum 8-hour ozone concentrations (in parts per million, ppm) were reported by 1,194 stations across the U.S. from January 1 to May 16, 2020. Daily average PM2.5 concentrations (in micrograms per cubic meter, μg/m^3^) were reported by 1,078 stations across the U.S. during the period. Each county-day with the ozone and PM2.5 information recorded by the first to the tenth nearest air quality monitors to the county’s centroid. More than one monitor’s recording was assigned to a county in order to estimate the nearest station’s missing values with the values recorded by the nearby stations.

The UV light index for the U.S. county centroids was obtained from the www.openweather.com, using the Python “pyowm” program,^57^ and joined to the data file. The assembled data file included COVID-19 cases and deaths, demographics, social distancing, weather, ozone, PM2.5, and UV information for each county-day from January 1 to May 16, 2020 (**Figure 1**).

### Data Refinement Procedure

The assembled data file included 2,984 counties. Because of missing values, only 41% of county-days were assigned with the temperature information from their nearest weather stations; 21%, 12%, 8%, and 6% of them were assigned with the temperature information from their second, third, fourth, and fifth nearest weather stations, respectively. The median distances of the first to fifth nearest stations from the county centroid were 4.5 (Standard Deviation, SD=5.5), 7.0 (SD=5.7), 9.9 (SD=6.7), 11.6 (SD=6.3), and 12.1 (SD=6.6) miles, respectively. There were fewer station changes in the assignment of precipitation, ozone, and PM2.5 information to counties than for pollution measures. At least 75% and 86% of county-days were assigned with the precipitation, ozone, and PM2.5 information from their first and first two nearest weather or air quality stations, respectively. The median distance of the nearest and the second nearest precipitation-recording station from the county centroid was 4.0 (SD=4.6) and 7.9 (SD=7.0) miles, respectively.

County-days that were assigned with the information from a weather station located 50 miles or more away from the county centroid were checked, and no such cases were identified. Among the county-days with missing weather information in the nearest station, checks were done to determine if weather station data were estimated with weather information from a station that was located 25 miles or more away from the nearest weather station, and these counties were excluded, resulting in 2,931 counties included in the analysis.

In general, the distances of ozone-and PM2.5-recording air quality stations from county centroids were longer than those of weather stations. In the unrefined data file, the median distances for the first and second nearest ozone-recording stations were 28.6 (SD=52.6) miles and 36.4 (SD=61.9) miles, respectively, and 30.6 (SD=35.6) and 40.9 (SD=34.2) for PM2.5. Such long distances can result in a potentially substantial error in the measurement of air pollutants at the county level. Thus, any county-day that was assigned with ozone or PM2.5 value recorded by a station that was located 60 miles or more away from the county centroid was dropped, and 594 counties were subsequently excluded from the analysis. Among the remaining 2,337 counties in the analysis, counties that had reported at least one COVID-19 death by May 16, 2020, were included to avoid measuring spurious effects. The latter constraint resulted in dropping 1,024 counties, leaving 1,313 counties in the sample. In the final work file for statistical analyses, another 30 counties were dropped due to the existence of missing values still remained in the data. Hence, the final work file included 1,273 counties from February 28 to May 16, 2020 (**Figure 2**).

**Figure 2.**
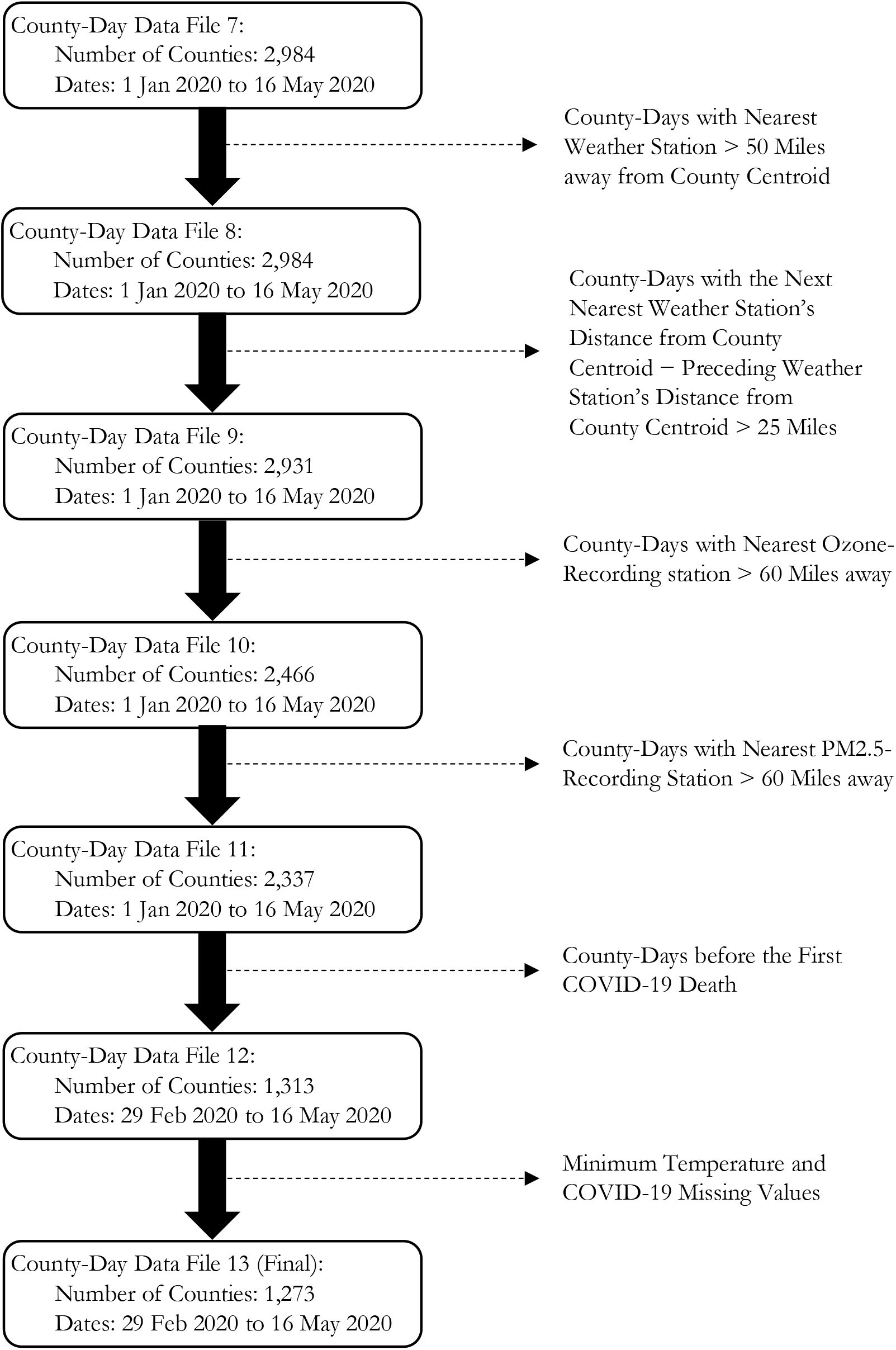
Data refinement procedure

The total number of county-days in the final work file was 42,674. In this final work file, the median distances of stations that recorded temperature, precipitation, ozone, and PM2.5 from the county centroid were 6.9 (SD=6.0), 4.2 (SD=4.8), 17.3 (SD=14.0), and 21.8 (SD=14.5) miles, respectively. The results of this study persisted lowering the maximum distance of stations that recoded to ozone and PM2.5 from 60 miles to 40 and 20 miles (**Table A1**). With the maximum 20 mile criterion, the median distances of stations that recorded ozone and PM2.5 were 9.5 (SD=5.4), and 10.5 (SD=5.7) miles from county centroids, respectively.

### Statistical Modeling

The logarithm of the daily COVID-19 deaths per capita in each county by day was the dependent variable in this analysis. Per capita was defined as the daily death rate by the size of the population over 18 years old in the county. The population below age 18 was excluded because COVID-19 rarely causes death in younger age cohorts. Daily death rate +1.0 was computed before dividing by population and taking the logarithm of the quotient to avoid an undefined term (i.e., the logarithm of zero). Four statistical models are described below. The first model was:

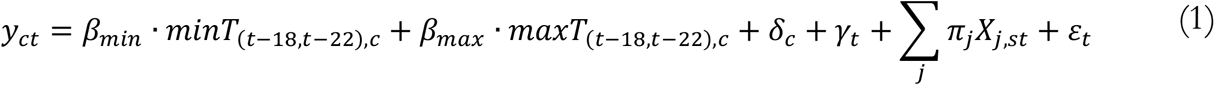

in which *y*_*ct*_is the log of daily death rate per capita in county *c* on day *t*. The published literature shows that, on average, infection occurs about 20 days before death.^58–63^ A five-day window around 20 days before death, which is 18 through 22 days before death, were taken and assumed that infection happened in that time window. In sensitivity analyses, this five-day window was changed to a three-day and a seven-day window and similar results were obtained (**Table A2-A5**). *minT*_(*t*−18,*t*−22),*c*_ and *maxT*_(*t*−18,*t*−22),*c*_ are the average minimum and maximum temperatures between 18 through 22 days before death (which his on day *t*). The coefficients of interest are *β*_*min*_ and *β*_*max*_.

*δ*_*c*_ is the set of county fixed-effects, where the subscript *c* represents a county. These are dummy variables for each county in the data. These fixed-effects control for factors that are constant over the period of this study but can be different across counties, and include factors such as demographic characteristics of a county, ethnic distribution in a county, health care features and resources of a county, cultural factors in a county, social norms, attitudes, and aspirations in a county, geography and climate of a county, political characteristics, and infrastructure of a county.

*γ*_*t*_ is a set of time fixed-effects for each day hence the subscript *t*. There were 78 days in the analysis sample. Hence, 78 dummy variables were included in the regression, each representing a day. These fixed-effects pick up any variable that is constant across counties in the U.S. but can vary daily. Federal government policies or anything that affects the whole nation (e.g., the Center for Disease Control’s measures, travel restrictions, and immigration policies) are examples of time fixed-effects.

Some policies and procedures in response to COVID-19 occurred at the state level in the U.S., vary by state, and are not the same across all counties in the U.S. within a given day. Hence, the day fixed-effects, *γ*_*t*_, cannot capture these potentially confounding factors. These policies were controlled using *X*_*j,st*_, which is a set of four policy-response dummies that control for four state-level policy changes over time: (1) shelter-in-place order, (2) order of no gathering of more than 500 people, (3) order of public school closures, and (4) order of closure of restaurants, entertainment venues, and gyms. In other words, for each policy, a dummy variable was defined that is equal to one for a county on a specific day if the state to which the county belongs had that policy in place on that day. The dummies are zero otherwise. Like temperature variables, the average of these dummy variables 18 through 22 days before death as independent variables.

To strengthen this model, Model (2) was defined below in which county-level time-trends, *θ*_*c*_*t*, were added:

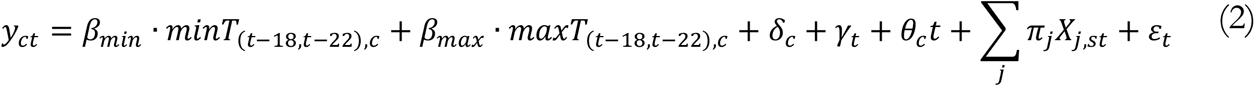

The time-trends control for the time trend in *y*_*ct*_(i.e., trends in the log of per capita daily death rate specific to each county). In other words, they allow each county to have its own time trend. Together, they constitute 1,453 additional independent variables in the regression.

In Models (1) and (2) include day fixed-effects, *γ*_*t*_, which control for any country-level factor such as federal-level policies that change daily but is constant across all counties on any day. Regional-level day fixed-effects were added to control for even more time-varying factors. These are day fixed-effects that are unique to a certain region of the U.S. That is, they control for factors that (1) can vary day by day, (2) are constant for counties within a certain region over any day, but (3) can be different for counties in other regions. For example, some neighboring states in certain regions coordinated their responses. Such coordination can mean that every day these states may implement similar policies that are unique to that region and period but different from other regions. The region-specific day fixed-effects, represented by *μ*_*rt*_ in Models (3) and (4), capture such factors. The regions used in this study were New England, Mid-East, Great Lakes, Plains, Southeast, Southwest, Rocky Mountain, and Far West.^64^

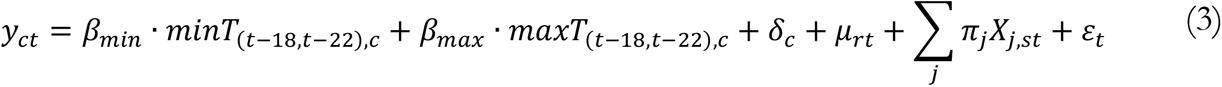

The number of dummy variables representing *μ*_*rt*_ fixed-effects in these models can be calculated as 8 regions *×*78 days = 624. In Model (4), county-level time-trends were added to Model (3):

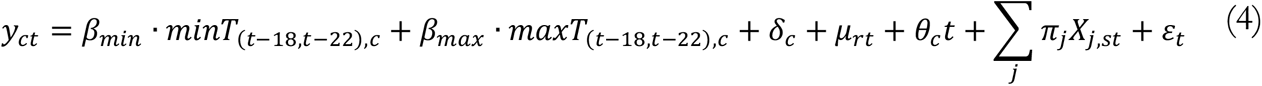

Model (4) provides the strongest specification and controls for numerous time-constant and time-varying factors. It includes 3,534 variables other than temperatures, some of which, like the county fixed-effects, or day fixed-effects control for multiple factors by themselves. This is the most robust model that has been used among studies related to weather and COVID-19 thus far.^20–41^ To strengthen the results, precipitation, pollution, and UV light were added as independent variables to Model (4) because these variables could be potential confounders. As with temperature variables, the averages of precipitation, pollution, and UV light, between 18 through 22 days before death (the most likely period of exposure to SARS-CoV-2) were added to the regressions.

Americans who live in the north of the U.S. may respond differently to changes in the temperature than those who live in the south due to the warmer climate. It is speculated that changes in temperature may change mobility and contact in the north differently from the south. Thus, death rates may vary differently with temperature in the north vs. the south U.S. To understand these differences, the sample was split into counties in the north and south, and the models were run separately for the north and south samples. The latitude of the geographical center of the continental United States, which is 39°50’ north,^65^ was used to split the sample. Counties with centroid latitudes greater than 39°50’ were included in the *North* sample, and those with centroid latitudes less than 39°50’ were included in the *South* sample.

One issue to be careful about is the serial or auto correlation. There is a robust solution to the problem of serial correlation in the panel data estimation: correcting the standard errors, hence p-values, of the coefficients by clustering them at the cross-sectional level. As a result, no lagged dependent variable will be needed as a regressor. In the present study, the cross-sectional units are counties. Thus, clustering standard errors at the county level accounts for the serially correlated structure of standard errors within each county.^66–69^ Thus, in all of the regressions for all models, p-values were clustered at the county level to address serial correlation.

## Results

There were 87,771 COVID-19 deaths in the U.S. by May 16, 2020.^70^ The study sample included 52,823 or 60% of these deaths. The majority, 70%, of the 52,823 deaths, occurred in the northern sample, the rest in the southern sample. On average, there were 1.2 (SD=5.8) deaths in the county-days of the analysis sample. The distribution of deaths was left-skewed because there were no COVID-19 deaths in 73% of the county-days and less than two deaths in 86% of county-days. The distribution of COVID-19 deaths over county-day was less left-skewed in the northern than the southern sample: in 67% and 77% of county-days, no COVID-19 deaths were reported in the north and south, respectively. Also, at least two COVID-19 deaths were reported in 18% and 9% of county-days in the north and south, respectively (**Table 1, Figure A1**). Because of the skewed distribution of the daily deaths, the logarithm of per capita deaths counts was used in the statistical analyses.

**Table 1.** Summary Statistics of COVID-19 death and weather and air quality indicators 18 to 22 days before death to COVID-19

| Variables | Number of<br>Counties | Number of<br>Observations | Mean | Standard<br>Deviation | Minimum | 25th<br>Quantile | Median | 75th<br>Quantile | 95th<br>Quantile | Maximum |
| --- | --- | --- | --- | --- | --- | --- | --- | --- | --- | --- |
| Full Sample |  |  |  |  |  |  |  |  |  |  |
| Number of New Deaths | 1,273 | 42,674 | 1.2 | 5.8 | 0.0 | 0.0 | 0.0 | 1.0 | 5.0 | 258.0 |
| log(Daily Deaths per 18+ Population) | 1,273 | 42,674 | 2.5 | 4.2 | 0.0 | 0.0 | 0.0 | 7.3 | 10.5 | 13.7 |
| Average Minimum Temperature (F) | 1,273 | 42,674 | 42.1 | 11.3 | -0.1 | 33.6 | 41.0 | 50.0 | 62.2 | 80.4 |
| Average Maximum Temperature (F) | 1,273 | 42,674 | 63.4 | 12.3 | 25.6 | 53.6 | 63.2 | 73.0 | 83.2 | 104.8 |
| Average Precipitation (mm) | 1,273 | 42,674 | 36.6 | 45.8 | 0.0 | 5.0 | 21.8 | 50.6 | 127.0 | 639.2 |
| Average Ozone Concentration (ppb) | 1,273 | 42,674 | 40.4 | 5.7 | 1.2 | 37.2 | 41.2 | 44.2 | 48.4 | 78.2 |
| Average PM2.5 Concentration ($\mu\text{g}/\text{m}^3$) | 1,273 | 42,674 | 7.3 | 3.1 | -0.4 | 5.2 | 6.8 | 8.9 | 12.6 | 37.1 |
| Average UV Light Index | 1,273 | 42,674 | 6.6 | 1.8 | 1.4 | 5.2 | 6.4 | 7.9 | 9.7 | 12.0 |
| Northern Sample |  |  |  |  |  |  |  |  |  |  |
| Number of New Deaths | 460 | 15,931 | 2.3 | 8.8 | 0.0 | 0.0 | 0.0 | 1.0 | 12.0 | 258.0 |
| log(Daily Deaths per 18+ Population) | 460 | 15,931 | 3.0 | 4.4 | 0.0 | 0.0 | 0.0 | 8.6 | 10.5 | 12.8 |
| Average Minimum Temperature (F) | 460 | 15,931 | 32.5 | 5.9 | -0.1 | 28.8 | 32.6 | 36.4 | 41.8 | 57.0 |
| Average Maximum Temperature (F) | 460 | 15,931 | 52.3 | 7.1 | 25.6 | 47.7 | 52.2 | 56.6 | 64.6 | 87.0 |
| Average Precipitation (mm) | 460 | 15,931 | 28.3 | 29.2 | 0.0 | 6.6 | 20.4 | 41.2 | 82.4 | 283.2 |
| Average Ozone Concentration (ppb) | 460 | 15,931 | 39.6 | 4.9 | 16.2 | 36.6 | 40.2 | 43.0 | 46.2 | 61.4 |
| Average PM2.5 Concentration ($\mu\text{g}/\text{m}^3$) | 460 | 15,931 | 6.5 | 2.4 | -0.4 | 4.7 | 6.3 | 7.9 | 11.0 | 17.7 |
| Average UV Light Index | 460 | 15,931 | 5.0 | 0.9 | 1.4 | 4.5 | 5.1 | 5.6 | 6.4 | 8.7 |
| Southern Sample |  |  |  |  |  |  |  |  |  |  |
| Number of New Deaths | 813 | 26,743 | 0.6 | 2.4 | 0.0 | 0.0 | 0.0 | 0.0 | 3.0 | 96.0 |
| log(Daily Deaths per 18+ Population) | 813 | 26,743 | 2.1 | 4.0 | 0.0 | 0.0 | 0.0 | 0.0 | 10.5 | 13.7 |
| Average Minimum Temperature (F) | 813 | 26,743 | 47.8 | 9.7 | 4.6 | 41.2 | 47.4 | 54.4 | 64.4 | 80.4 |
| Average Maximum Temperature (F) | 813 | 26,743 | 70.0 | 9.7 | 26.8 | 63.4 | 70.2 | 77.2 | 85.0 | 104.8 |
| Average Precipitation (mm) | 813 | 26,743 | 41.6 | 52.7 | 0.0 | 3.6 | 22.8 | 59.4 | 150.0 | 639.2 |
| Average Ozone Concentration (ppb) | 813 | 26,743 | 40.9 | 6.2 | 1.2 | 37.6 | 41.8 | 45.0 | 49.4 | 78.2 |
| Average PM2.5 Concentration ($\mu\text{g}/\text{m}^3$) | 813 | 26,743 | 7.7 | 3.4 | -0.1 | 5.5 | 7.3 | 9.4 | 13.6 | 37.1 |
| Average UV Light Index | 813 | 26,743 | 7.5 | 1.5 | 2.9 | 6.5 | 7.5 | 8.6 | 10.1 | 12.0 |

Notes: Numbers in parentheses are standard deviations. Counties whose centroid is above the 39° 50'N are considered northern. Temperatures are in Fahrenheit. Precipitation is in millimeters. Ozone is the maximum 8-hour concentration level measured in parts per billion, ppb. PM2.5 is the average daily concentration level measured in micrograms per cubic meters, $\mu\text{g}/\text{m}^3$ .

The average minimum temperature during the presumed days of exposure to the coronavirus (18 through 22 days before death) was 42.1°F (SD=11.3°F). It was lower in the northern sample, 32.5°F (SD=5.9°F), but higher in the southern sample, 47.8°F (SD=9.7°F). The average maximum temperature during the exposure period was 63.4°F (SD=12.3°F), with a northern-southern gradient of 52.3°F (SD=7.1°F) and 70.0°F (SD=9.7°F), respectively. On average, there were 36.6 mm (SD=45.8 mm) of rainfall during the exposure period, 28.3 mm (SD=29.2 mm) in the northern and 41.6 mm (SD=52.7 mm) in the southern samples, respectively. The three meteorological elements were approximately normally distributed, except precipitation. There was also greater variation in precipitation than temperatures across counties over time. The logarithm of precipitation (plus one) was obtained to turn it into a distribution that resembles a normal distribution (**Table 1, Figure A2**).

The average of 8-hour maximum concentration of the ground-level ozone during the exposure period was 40.4 ppb (SD=5.7 ppb). While the average ozone levels were generally in the “Good” AQI (air quality index) range,^71^ the median ozone concentration level in the northern U.S. was sless than that in the southern U.S., 40.2 ppb (SD=4.9 ppb) versus 41.8 ppb (SD=6.2 ppb). The highest ozone level in the south was 78.2 ppb, categorized as “Unhealthy for Sensitive Groups” by the EPA,^71^ but 61.4 ppb, categorized as “Moderate,” in the north. In addition, there was more variation in ozone in the south (**Table 1, Figure A2**). The averages of the mean daily PM2.5 concentration and the UV light index in the analysis sample were 7.3 μg/m^3^ (SD=3.1 μg/m^3^) and 6.6 (SD=1.8), respectively. Again, the average, variation, and maximum level of both PM2.5 and UV light index were greater in the south (**Table 1, Figure A2**).

The association of COVID-19 deaths at the county-level in the U.S. was estimated with minimum and maximum daily temperatures during the exposure period, while systematically adding control variables to assess the effect of each potential contributing factor on the association (**Table 2**). As basic control variables, the following were included in all model specifications: four social distancing measures (shelter-in-place, no large gathering, closure of public schools, and closure of restaurants and entertainment venues), county fixed-effects, and day fixed-effects. The latter set of controls varied in some specifications. The estimates from the first model, which include U.S.-level day fixed-effects, show that a 1°F increase in the five-day average of the daily minimum temperature was associated with a 3.8% increase in adult COVID-19 deaths in a typical U.S. county during the study period. County-level daily death rates in the left-hand-side (dependent variable) of all statistical models were weighted by county population 18 years and older. Thus, the estimated association can also be interpreted as the increase in the number of COVID-19 deaths per 18 years and older county population. Note that a 1°F increase in the five-day average of daily minimum temperature requires the daily minimum temperature to increase by 5°F cumulatively in those five days. This is not necessarily a small change. Also, a 1°F increase in the five-day average of the daily maximum temperature was associated with a 2.1% decrease in adult COVID-19 deaths per 18 years and older county population during the study period.

**Table 2.** Multiple regression results of estimating the association of weather and air quality indicators with the logarithm of county-level daily death rate 18+ population in 1,273 counties and 42,674 county days *Interpretation of the reported coefficients:* Percentage (in decimals) change in daily death per 18+ county population for a one-unit change in the weather and air quality indicator.

| Variables | (1)<br>Model 1 | (2)<br>Model 2 | (3)<br>Model 3 | (4)<br>Model 4 | (5)<br>Model 4 | (6)<br>Model 4 | (7)<br>Model 4 | (8)<br>Model 4 |
| --- | --- | --- | --- | --- | --- | --- | --- | --- |
| Average Minimum Temperature (F) | 0.038*** | 0.026*** | 0.026*** | 0.024*** | 0.025*** | 0.020** | 0.020** | 0.018* |
| p-value | (0.00) | (0.00) | (0.00) | (0.01) | (0.01) | (0.04) | (0.04) | (0.06) |
| Average Maximum Temperature (F) | -0.021*** | -0.011** | -0.004 | 0.001 | 0.000 | 0.005 | 0.005 | 0.004 |
| p-value | (0.00) | (0.05) | (0.52) | (0.86) | (0.97) | (0.48) | (0.50) | (0.60) |
| Log(Average Precipitation (mm)) |  |  |  |  | -0.008 | -0.013 | -0.011 | -0.010 |
| p-value |  |  |  |  | (0.67) | (0.52) | (0.57) | (0.62) |
| Average Ozone Concentration (ppb) |  |  |  |  |  | -0.014* | -0.014** | -0.013* |
| p-value |  |  |  |  |  | (0.05) | (0.05) | (0.07) |
| Average PM2.5 Concentration ($\mu\text{g}/\text{m}^3$) | | | | | | | 0.005 | 0.004 |
| p-value |  |  |  |  |  |  | (0.71) | (0.76) |
| Average UV Light Index |  |  |  |  |  |  |  | 0.128 |
| p-value |  |  |  |  |  |  |  | (0.25) |
| Set of Geographical and Time Controls: |  |  |  |  |  |  |  |  |
| Social Distancing Measures | Yes | Yes | Yes | Yes | Yes | Yes | Yes | Yes |
| County Fixed-Effects | Yes | Yes | Yes | Yes | Yes | Yes | Yes | Yes |
| US-Level Day Fixed-Effects | Yes | Yes |  |  |  |  |  |  |
| Region-Level Day Fixed-Effects |  |  | Yes | Yes | Yes | Yes | Yes | Yes |
| County-Specific Time-Trends |  | Yes |  | Yes | Yes | Yes | Yes | Yes |
| R-squared | 0.021 | 0.095 | 0.043 | 0.110 | 0.109 | 0.109 | 0.109 | 0.109 |

*Notes:* All standard errors are clustered at the county level. Numbers in parentheses are robust p-values. Signs \*\*\*, \*\*, and \* indicate 1%, 5%, and 10% statistical significance levels. The minimum and maximum temperatures are in Fahrenheit. Precipitation is in millimeters. Ozone is the maximum 8-hour concentration level measured in parts per billion, ppb. PM2.5 is the average daily concentration level measured in micrograms per cubic meters, $\mu\text{g}/\text{m}^3$ .

The negative association of COVID-19 deaths with the five-day average maximum temperature became non-significant when the model was improved by including county-specific time trends. Also, the size of the positive association of COVID-19 deaths with the five-day average minimum temperature decreased. The estimates from the second model show that a 1°F increase in the five-day average minimum temperature was associated with a 2.6% increase in county-level COVID-19 deaths during the study period (**Table 2**). The results from Model (2) largely sustained the replacement of the U.S.-level fixed-effects with the region-level fixed-effects and the dropping of county-specific time trends, Models (3) and (4). According to the estimates from the specification with the preferred set of time-constant and time-varying controls but without other weather elements and air pollutants, Model (4), a 1°F increase in the five-day minimum temperature was associated with a 2.4% increase in county-level COVID-19 deaths during the study period (**Table 2**).

Model (4) was improved by adding daily precipitation, ozone, PM2.5, and UV index. The factor with the greatest influence on the association of county-level COVID-19 deaths with daily minimum temperature was ozone concentration, which resulted in a 20% decrease in the magnitude of the association (**Table 2**). Ozone itself also had a statistically significant negative association with COVID-19 deaths.

According to the estimates from the optimal statistical model, Model (4), a 1°F increase in the five-day average minimum temperature was associated with a 1.8% increase in county-level COVID-19 deaths during the study period. The association between COVID-19 deaths in a county and the daily maximum temperature, daily precipitation, PM2.5 concentration, and UV light index was not statistically significant. However, a 1 ppb increase in the 8-hour maximum ozone concentration level during the day was associated with a 1.3% decrease in county-level COVID-19 deaths during the study period (**Table 2**).

The associations of COVID-19 deaths with minimum daily temperature and ozone differed between the U.S. north and south regions. Specifically, the positive association between COVID-19 deaths and the minimum daily temperature was statistically significant only in the northern sample during the study period, but no such association was detected in the southern sample. In the north, a 1°F increase in the five-day average of the minimum daily temperature was associated with a 5.1% increase in county-level COVID-19 deaths during the study period. No other statistically significant associations were detected in the analysis of the northern sample (**Table 3**). To put this result in context, suppose that a northern county experienced 100 COVID-19 deaths on a day (for example, April 23) and suppose that on every day between 18 through 22 days before that day (i.e., April 1-5) the minimum temperature was X degrees. Thus, the five-day average minimum temperature was also equal to X degrees. On the following day (April 24), approximately 105 people will die of COVID-19 if the five-day average minimum temperature for that day increased by 1°F (i.e., the average minimum temperature for April 2-6). The minimum temperature should have increased by 5°F on April 6 so that the five-day average minimum temperature for April 24 increased by 1°F.

**Table 3.** Multiple regression results of estimating the association of weather and air quality indicators with the logarithm of county-level daily deaths per 18+ population in northern and southern counties, using the preferred statistical model (Model 4). *Interpretation of the reported coefficients:* Percentage (in decimals) change in daily death per 18+ county population for a one-unit change in the weather and air quality indicator.

| Variables | Per 18+ County Population |  |  |
| --- | --- | --- | --- |
|  | Full Sample | Northern Sample | Southern Sample |
| Average Minimum Temperature (F) | 0.018* | 0.051*** | -0.005 |
| p-value | (0.06) | (0.01) | (0.68) |
| Average Maximum Temperature (F) | 0.004 | -0.002 | 0.004 |
| p-value | (0.60) | (0.90) | (0.69) |
| Log(Average Precipitation (mm)) | -0.010 | 0.043 | -0.027 |
| p-value | (0.62) | (0.20) | (0.29) |
| Average Ozone Concentration (ppb) | -0.013* | 0.007 | -0.020** |
| p-value | (0.07) | (0.62) | (0.02) |
| Average PM2.5 Concentration ($\mu\text{g}/\text{m}^3$) | 0.004 | 0.028 | 0.004 |
| p-value | (0.76) | (0.35) | (0.80) |
| Average UV Light Index | 0.128 | -0.113 | 0.096 |
| p-value | (0.25) | (0.72) | (0.46) |
| Set of Geographical and Time Controls: |  |  |  |
| Social Distancing Measures | Yes | Yes | Yes |
| County Fixed-Effect | Yes | Yes | Yes |
| Region-Level Day Fixed-Effect | Yes | Yes | Yes |
| County Time-trends | Yes | Yes | Yes |
| Number of Counties | 1,273 | 460 | 813 |
| Observations | 42,674 | 15,931 | 26,743 |
| R-squared | 0.114 | 0.151 | 0.109 |

*Notes:* Counties whose centroid is above the 39°50'N are considered northern. All standard errors are clustered at the county level. Numbers in parentheses are robust p-values. Signs \*\*\*, \*\*, and \* indicate 1%, 5%, and 10% statistical significance levels. The minimum and maximum temperatures are in Fahrenheit. Precipitation is in millimeters. Ozone is the maximum 8-hour concentration level measured in parts per billion, ppb. PM2.5 is the average daily concentration level measured in micrograms per cubic meters, $\mu\text{g}/\text{m}^3$ .

In contrast, the negative association between COVID-19 deaths and the maximum 8-hour ozone concentration was statistically significant only in the southern sample, while no such association was detected in the northern sample. In the south, a 1 ppb increase in the five-day average ozone concentration was associated with a 2.0% decrease in county-level COVID-19 deaths during the study period. No other statistically significant associations were detected in the analysis of the southern sample (**Table 3**). To put this result in context, suppose that a southern county experienced 100 COVID-19 deaths on a day (for example, April 23) and suppose that on every day between 18 through 22 days before that day (i.e., April 1-5) the ozone concentration was Z ppb. Thus, the five-day average ozone concentration was also equal to Z ppb. On the next day (April 24), approximately 98 new COVID-19 deaths will be recorded if the five-day average ozone concentration for that day increased by 1 ppb (i.e., average ozone concentration for April 2-6). The ozone concentration should have increased by 5 ppb on April 6, so that the five-day average minimum temperature increased by 1 ppb.

## Discussion

### Interpretation of The Results

The average minimum daily temperature in the northern sample was 32.2°F. In 95% of county-days in the north, the minimum daily temperature was less than 42.2°F (**Table 1**). This range of minimum daily temperatures is below the range of temperatures in which SARS-CoV-2’s persistence on surfaces and in the aerosols decreases at elevated speeds.^14,72^ It is possible that the measured positive association between COVID-19 deaths and daily minimum temperature in the north is attributable to changes in contact rates (after accounting for the county, state, and time-specific contributing factors) because a likely increase in the minimum daily temperature was not likely to kill SARS-CoV-2 on surfaces or in the air. Warmer temperature may significantly increase the contact rate, which affects opportunities for the transmission of the virus. In the south, where no significant association between the five-day average minimum daily temperature and COVID-19 death was found, the increase in the minimum temperature during the period apparently did not significantly change contact rate.

An association between COVID-19 deaths and the maximum daily temperature is potentially the result of two opposing factors. The five-day average maximum daily temperatures were large enough that a substantial increase was likely to decrease the SARS-CoV-2 transmission rate. At the same time, temperature may increase the person-to-person contact rate. If no association is detected, it is possible that these two opposing forces balance out such that the impact of the increase in the five-day maximum daily temperature during the study period is not identified. Alternatively, it may indicate that the increases in the maximum daily temperature during the study period, which covers late winter and spring, were not sufficiently strong to have a disinfection effect against SARS-CoV-2 and did not change contact rates either.

The negative association between the airborne ozone level and COVID-19 deaths is a potentially important finding. Municipal sanitizing systems often use ozone to disinfect water sources or in some health care settings to disinfect surfaces. Laboratory testing shows that ozone may inactivate SARS-CoV-2.^55,56^ Currently, researchers are exploring an ozone-based intervention to curb the progression of CVOD-19.^73^ Also, there was a negative, although uncontrolled, correlation between average ozone level in major Chinese cities in January and March 2020 and confirmed COVID-19 cases.^74^ A global study also showed the negative association between COVID-19 transmission rate and ozone concentration.^26^ This study provides the first empirically robust evidence of a relationship between ozone and COVID-19 deaths.

### Comments on Methodology

The outcome analyzed is the COVID-19 death rate by day by county. The number of infected cases is limited by testing capacity and is not a random sample. In the U.S., like most countries, testing capacity in the period of this study was confounded by a proportionately small number of tests done for prevalence estimates, and far below the level to cover everyone. Although this capacity has increased over time, available data on the number of cases suffers substantially from a time-varying measurement error that may not be captured by often-used time variables, such as time trends. Some related studies that use the number of cases argued that as long as the measurement error in the number of cases is not correlated with daily changes in temperature, the results are reliable.^26^ This assumption, however, is unverifiable. The number of COVID-19 deaths, on the other hand, is more reliable than the number of cases because it has been collected with more precision. A potential measurement error in the number of COVID-19 deaths is the underdiagnosis of the disease in some fatalities and symptomologic diagnosis in others.

The focus of the present study is on the United States because the quality of data is more homogenous within a nation than between them, even for death. Reporting systems are likely to be more consistent within a given nation than across a number of countries. Therefore, U.S. data was the most consistent and reliable data on COVID-19 deaths that were accessible at the county level, and there are hundreds of counties in the analysis sample of this study.

Since there are 1,273 counties in the analysis sample, *δ*_*c*_ in Models (1)-(4) represents a collection of 1,273 dummy variables, known as fixed-effects. These county fixed-effects control for any confounder that is constant over the period of this study but can vary across counties. One benefit of county fixed-effects is that they control for any time-constant factor in a county even if the data on the factor is not available or the factor is not measurable. For example, if warmer counties have different health care infrastructure than cooler counties in general, county fixed-effects control for them as long as the health care infrastructure remains the same over the period of this study in each of these counties.

The day fixed-effects capture factors affecting the daily death rate that varies daily but is constant across counties in the United States. Federal government responses to COVID-19, CDC measures, or anything that affects the whole nation and the spread of COVID-19 (e.g., travel restrictions and immigration restrictions). They affect all counties, but their intensity or nature can vary from one day to another. It is necessary to control for time in a granular way without restriction on the functional form because the policy response to COVID-19 was ever-evolving during the period of this study. Controlling for a polynomial functional form of time trends does not capture all the variation in policy over time and is restricted by the functional form. Day fixed-effects, however, capture any non-linear changes in policies as they have no restriction on the functional form.

Separate day fixed-effects were defined for each region to strengthen day fixed-effects, in Model (4). This captures not only federal-level responses that vary daily, but also responses specific to each of the eight U.S. regions that can vary daily. This strong specification, together with controlling for state-specific policy measures (*X*_*j,st*_), captures numerous time-varying factors that can confound the results. In addition to these day fixed-effects, county time trends allow control for any trend in death rate within each county separately. This captures any trend in COVID deaths within a county that could be for any reason, such as the gradual increase in the understanding of the county population about the disease and ways to prevent it.

Because fixed-effects capture all independent variables that only vary by either county or time, independent variables added to these models must vary by both county and time. In addition, such independent variables should be potentially correlated with COVID death and temperature, and would need to be included in the model. Precipitation, pollution, and UV level are such potential variables. All these variables are included in this study’s models, making them powerful in the identification of the causal impact of temperature variation on COVID-19 deaths.

An important advantage of this study is that it addresses serial correlation in the error terms of the statistical models. A robust solution to the problem of serial correlation in this panel data estimation is to correct the standard errors and p-values of the coefficients by clustering them at the geographic level, which is the county. Thus, in all of the regressions for all models, reported p-values are corrected by clustering at the county level. This is an important issue that is often overlooked in other studies on the effect of environmental factors on COVID.

The R-squared of the models of this study is generally low. This is certainly not a disadvantage of these models. To identify the causal effects of an independent variable on a dependent variable, in regular statistical models, researchers include as many covariates as possible to capture the effect of confounders. Increasing the number of covariates in such models increases R-squared. Hence, one quick way for researchers to find whether confounders are sufficiently controlled for has been to examine the R-square values of such models. Fixed-effect models, by nature, produce small within-group R-squared because they control for an immense number of confounders, many of which are impossible or hard to measure.

### Limitations

The present study has several important limitations. Measurement error in the assignment of air pollution information is much greater than the error in the assignment of temperature and precipitation information. Specifically, the median distance of stations reporting temperatures, precipitation, ozone, and PM2.5 to the county centroid was 6.9, 4.2, 17.3, and 21.8 miles, respectively. The sensitivity of this study’s results was tested against the exclusion of counties with air quality stations 40 miles or more and 20 miles or more away from the county centroid. The results sustained the test. The association of COVID-19 deaths and the five-day average minimum temperature was 5.5% when the most restricted sample was used. The association of COVID-19 deaths and the five-day average ozone level, however, increased from −2.0% to −3.8% (**Table A1**). Moreover, the results were preserved after distances from weather and air quality stations were added as covariates to the statistical models (**Tables A6** and **A7**).

Another issue of note is that state-level day fixed-effects were not included in the models. Including state-level day fixed-effects removes much of the daily variation in temperature because many U.S. States, particularly in the Northeast, are too small to have much within-state variation in temperature. In other words, the results would produce a statistically insignificant estimate due to the model, rather than the true coefficient (A recently published study used the same set of region-level time fixed-effects as Model 3 to estimate the effect of social distancing policy measures on the spread of the virus.^52^). Nevertheless, a model with state-level day fixed-effects was estimated to compare the results:

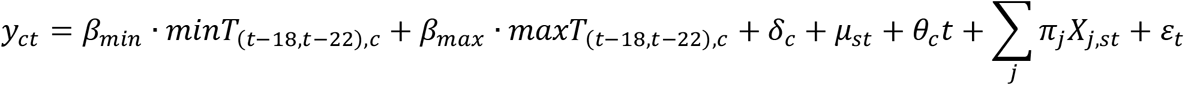

In this model, *μ*_*st*_ is a set of dummy variables representing state *s* and day *t*. They control for any factor that can vary daily within states. Interestingly, the results for the minimum temperature, reported in **Table A8**, remained the same for the northern sample. The results for the ozone concentration in the southern sample, however, faded away. As mentioned, this model controls for too many confounders and diminishes the variation in independent variables.

## Conclusion

Temperature and pollution are weather factors that could affect the spread of SARS-CoV-2. This study used the most reliable data, deaths, and a strong statistical model at the county level to estimate the impact of these factors. During March, April, and May of 2020, when the average minimum temperature within a five-day window increased by one degree Fahrenheit, daily death in northern U.S. counties increased by about 5.1%. In addition, when ozone concentration rised by one ppb, daily death in southern U.S. counties increases by about 2.0%. No evidence of an effect of maximum temperature, precipitation, PM2.5, and UV light on COVID-19 deaths was found. The absence of evidence, however, is not evidence of absence.

These results were robust to the specification and to the inclusion and exclusion of variables. The impact of minimum temperature on COVID-19 deaths is thought to be through the increase in physical mobility and personal contacts. Importantly, these results apply only to the spring weather during March, April, and May of 2020. The rise in the minimum temperature in the springtime in the northern United States could be the reason people leave their homes more often than in colder temperatures early in the year. Temperature (minimum) may not have the same effect on behavior in summer, but that was not yet analyzed. The impact of summer (or fall) weather on COVID-19 deaths will be separately analyzed. The effect of ozone may be explained by its disinfectant properties, but this has not been analyzed in laboratory experiments to our knowledge.

## Data Availability

All the data used in this study are publicly available. Data sources and the related links are provided in the text.

## Appendix Figures

**Figure A1.**
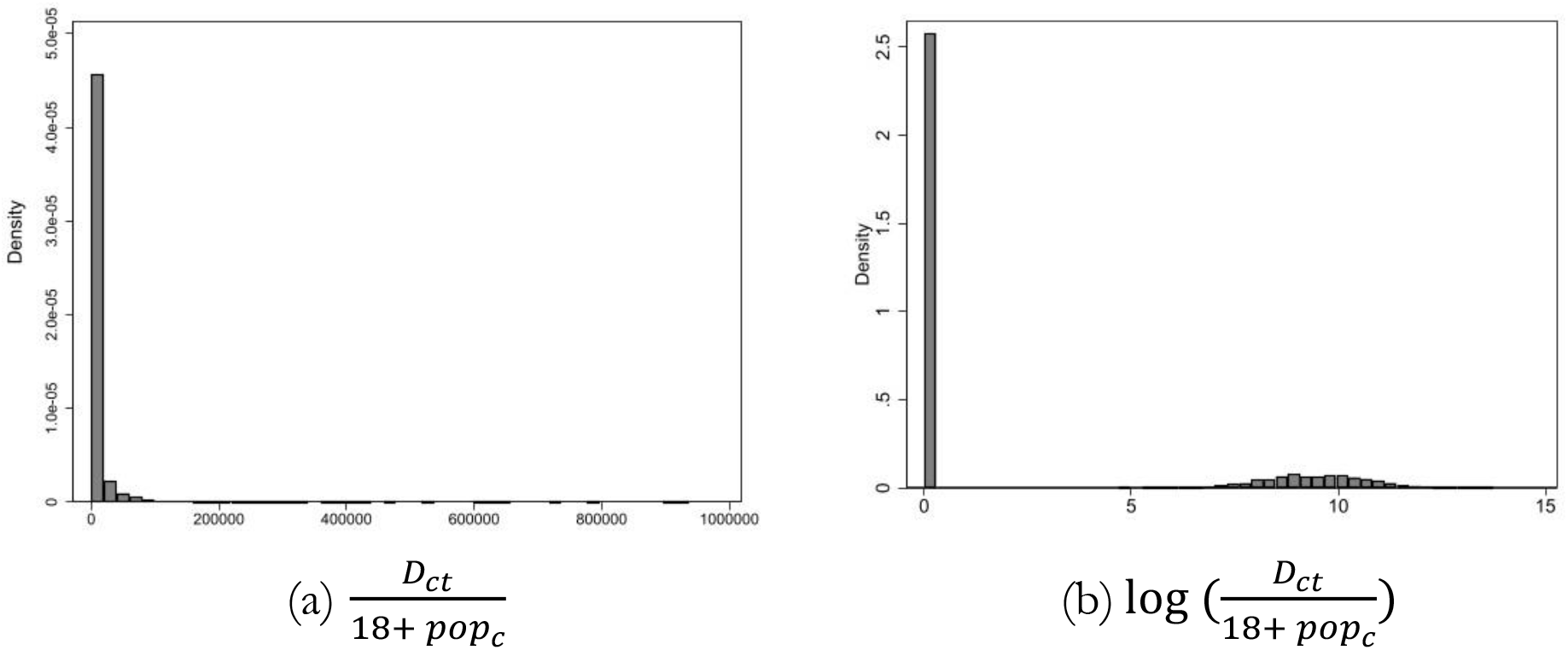
Histogram of Daily Death Rates. Note: *D*_*ct*_ stands for the number of new deaths in county *c* on day *t*. 18+ *pop*_*c*_ is the size of the population over age 18. Figure (a) shows the daily death rate per 18+ population in county *c* on day *t*. Figure (b) is the logarithm of this measure, used as the dependent variable in this study.

**Figure A2.**
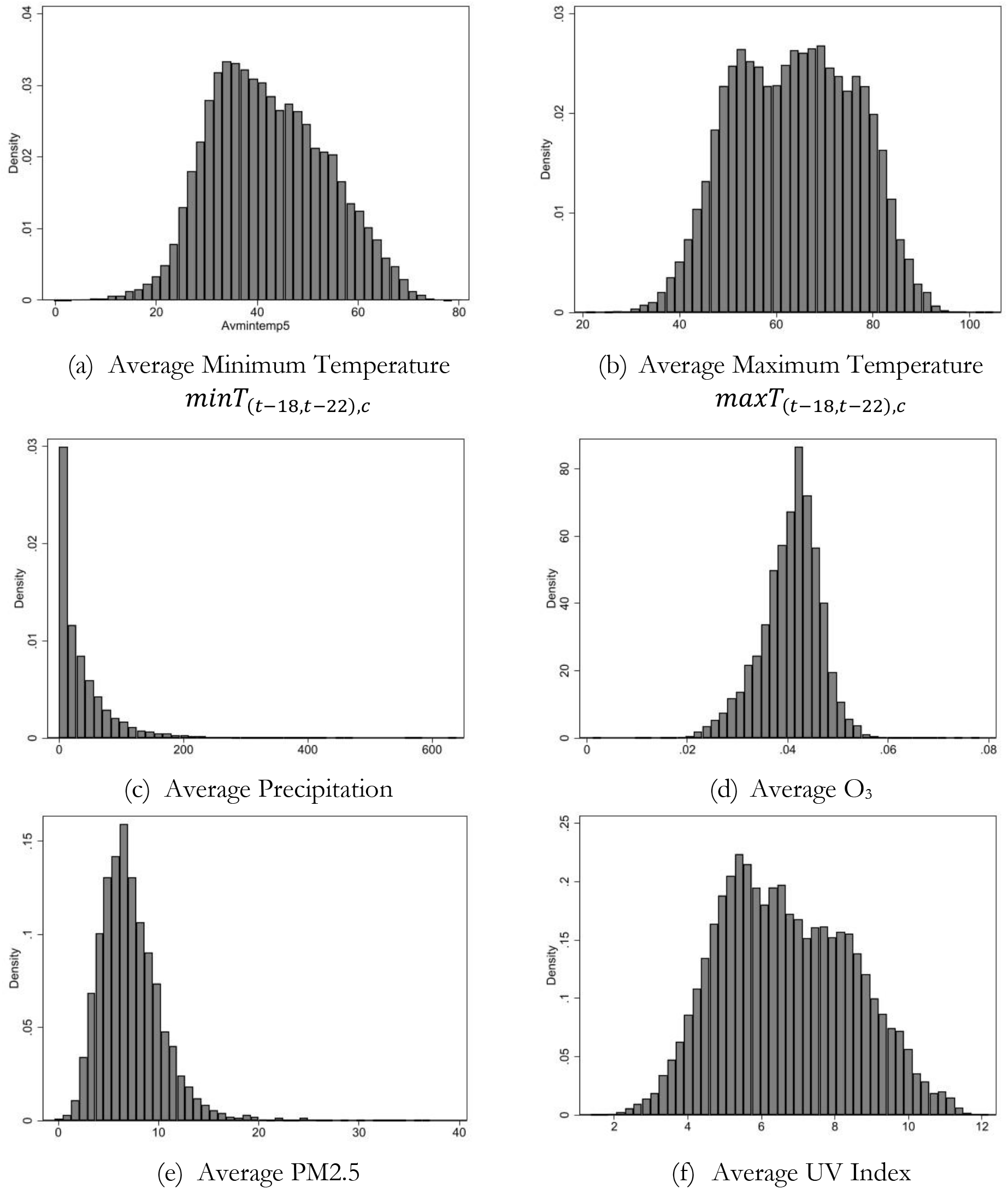
Histograms of Weather Variables. Note: All these averages are calculated over a five-day window 18 through 22 days before death.

## Appendix Tables

**Table A1.** Multiple regression results of estimating the association of weather and air quality indicators with the logarithm of county-level daily death rates (per 18+ population) for the northern and southern samples – Samples based on the distance of stations reporting ozone and PM data to the county centroid *Interpretation of the reported coefficients:* Percentage (in decimals) change in daily death per 18+ county population for a one-unit change in the weather and air quality indicator.

| Variables | Northern Sample |  |  |  |  | Southern Sample |  |  |  |  |
| --- | --- | --- | --- | --- | --- | --- | --- | --- | --- | --- |
|  | 20 miles | 40 miles | 60 miles | 80 miles | 100 miles | 20 miles | 40 miles | 60 miles | 80 miles | 100 miles |
| Average Minimum Temperature (F) | 0.055* | 0.055*** | 0.051*** | 0.042** | 0.038** | -0.022 | -0.015 | -0.005 | 0.000 | -0.001 |
| p-value | (0.06) | (0.01) | (0.01) | (0.01) | (0.02) | (0.30) | (0.25) | (0.68) | (0.97) | (0.95) |
| Average Maximum Temperature (F) | 0.012 | 0.003 | -0.002 | 0.001 | 0.004 | 0.025 | 0.008 | 0.004 | 0.001 | 0.005 |
| p-value | (0.58) | (0.87) | (0.90) | (0.94) | (0.74) | (0.17) | (0.52) | (0.69) | (0.91) | (0.57) |
| Log (Average Precipitation (mm)) | 0.027 | 0.052 | 0.043 | 0.037 | 0.030 | -0.077* | -0.021 | -0.027 | -0.027 | -0.024 |
| p-value | (0.60) | (0.18) | (0.20) | (0.23) | (0.33) | (0.08) | (0.49) | (0.29) | (0.24) | (0.29) |
| Average O<sub>3</sub> Concentration (ppb) | 0.003 | 0.005 | 0.007 | 0.012 | 0.010 | -0.038** | -0.029*** | -0.020** | -0.021*** | -0.017** |
| p-value | (0.89) | (0.77) | (0.62) | (0.38) | (0.48) | (0.02) | (0.00) | (0.02) | (0.01) | (0.02) |
| Average PM<sub>2.5</sub> Concentration (µg/m<sup>3</sup>) | 0.003 | 0.023 | 0.028 | 0.021 | 0.024 | -0.046* | -0.005 | 0.004 | -0.004 | -0.006 |
| p-value | (0.95) | (0.51) | (0.35) | (0.44) | (0.37) | (0.07) | (0.76) | (0.80) | (0.78) | (0.63) |
| Average UV Light Index | -0.173 | -0.234 | -0.113 | -0.060 | 0.036 | 0.082 | 0.242* | 0.096 | 0.046 | 0.021 |
| p-value | (0.75) | (0.53) | (0.72) | (0.83) | (0.90) | (0.68) | (0.09) | (0.46) | (0.71) | (0.86) |
| Set of Geographic and Time Controls: |  |  |  |  |  |  |  |  |  |  |
| Social Distancing Measures | Yes | Yes | Yes | Yes | Yes | Yes | Yes | Yes | Yes | Yes |
| County Fixed-Effect | Yes | Yes | Yes | Yes | Yes | Yes | Yes | Yes | Yes | Yes |
| Region-Level Day Fixed-Effect | Yes | Yes | Yes | Yes | Yes | Yes | Yes | Yes | Yes | Yes |
| County Time-trends | Yes | Yes | Yes | Yes | Yes | Yes | Yes | Yes | Yes | Yes |
| Number of Counties | 184 | 379 | 460 | 488 | 501 | 274 | 628 | 813 | 896 | 934 |
| Observations | 6,731 | 13,101 | 15,931 | 16,947 | 17,559 | 9,573 | 20,613 | 26,743 | 29,882 | 31,291 |
| R-squared | 0.224 | 0.161 | 0.151 | 0.147 | 0.145 | 0.152 | 0.117 | 0.109 | 0.106 | 0.105 |

*Notes:* All standard errors are clustered at the county level. Numbers in parentheses are robust p-values. Signs \*\*\*, \*\*, and \* indicate 1%, 5%, and 10% statistical significance levels. The minimum and maximum temperatures are in Fahrenheit. Precipitation is in millimeters. O<sub>3</sub> is the maximum 8-hour concentration level measured in parts per billion, ppb. PM<sub>2.5</sub> is the average daily concentration level measured in micrograms per cubic meters, µg/m<sup>3</sup>.

**Table A2.** Multiple regression results of estimating the association of weather and air quality indicators with the logarithm of county-level daily death rate per 18+ population in 1,261 counties and 41,958 county days - Average weather variables are calculated over a 7-day period: 17 through 23 days before death. *Interpretation of the reported coefficients:* Percentage (in decimals) change in daily death per 18+ county population for a one-unit change in the weather and air quality indicator.

| Variables | (1)<br>Model 1 | (2)<br>Model 2 | (3)<br>Model 3 | (4)<br>Model 4 | (5)<br>Model 4 | (6)<br>Model 4 | (7)<br>Model 4 | (8)<br>Model 4 |
| --- | --- | --- | --- | --- | --- | --- | --- | --- |
| Average Minimum Temperature (F) | 0.041*** | 0.027*** | 0.028** | 0.029** | 0.031*** | 0.019 | 0.019 | 0.018 |
| p-value | (0.00) | (0.00) | (0.01) | (0.01) | (0.01) | (0.11) | (0.12) | (0.15) |
| Average Maximum Temperature (F) | -0.022*** | -0.008 | -0.003 | 0.004 | 0.000 | 0.012 | 0.012 | 0.010 |
| p-value | (0.00) | (0.27) | (0.71) | (0.65) | (0.98) | (0.25) | (0.27) | (0.33) |
| Log (Average Precipitation (mm)) |  |  |  |  | -0.033 | -0.041* | -0.038 | -0.037 |
| p-value |  |  |  |  | (0.17) | (0.09) | (0.13) | (0.14) |
| Average O<sub>3</sub> Concentration (ppb) |  |  |  |  |  | -0.030*** | -0.030*** | -0.028*** |
| p-value |  |  |  |  |  | (0.00) | (0.00) | (0.00) |
| Average PM<sub>2.5</sub> Concentration (µg/m<sup>3</sup>) |  |  |  |  |  |  | 0.010 | 0.009 |
| p-value |  |  |  |  |  |  | (0.57) | (0.62) |
| Average UV Light Index |  |  |  |  |  |  |  | 0.161 |
| p-value |  |  |  |  |  |  |  | (0.22) |
| Set of Geographic and Time Controls: |  |  |  |  |  |  |  |  |
| Social Distancing Measures | Yes | Yes | Yes | Yes | Yes | Yes | Yes | Yes |
| County Fixed-Effects | Yes | Yes | Yes | Yes | Yes | Yes | Yes | Yes |
| US-Level Day Fixed-Effects | Yes | Yes |  |  |  |  |  |  |
| Region-Level Day Fixed-Effects |  |  | Yes | Yes | Yes | Yes | Yes | Yes |
| County-Specific Time-Trends |  | Yes |  | Yes | Yes | Yes | Yes | Yes |
| R-squared | 0.023 | 0.097 | 0.046 | 0.114 | 0.114 | 0.114 | 0.114 | 0.114 |

*Notes:* All standard errors are clustered at the county level. Numbers in parentheses are robust p-values. Signs \*\*\*, \*\*, and \* indicate 1%, 5%, and 10% statistical significance levels. The minimum and maximum temperatures are in Fahrenheit. Precipitation is in millimeters. O<sub>3</sub> is the maximum 8-hour concentration level measured in parts per billion, ppb. PM<sub>2.5</sub> is the average daily concentration level measured in micrograms per cubic meters, µg/m<sup>3</sup>.

**Table A3.** Multiple regression results of estimating the association of weather and air quality indicators with the logarithm of county-level daily deaths per 18+ population in northern and southern counties, using the preferred statistical model (Model 4) - Average weather variables are calculated over a 7-day period: 17 through 23 days before death. *Interpretation of the reported coefficients:* Percentage (in decimals) change in daily death per 18+ county population for a one-unit change in the weather and air quality indicator.

| Variables | Per 18+ County Population |  |  |
| --- | --- | --- | --- |
|  | Full Sample | Northern Sample | Southern Sample |
| Average Minimum Temperature (F) | 0.018 | 0.082*** | -0.024 |
| p-value | (0.15) | (0.00) | (0.10) |
| Average Maximum Temperature (F) | 0.010 | -0.014 | 0.025* |
| p-value | (0.33) | (0.46) | (0.07) |
| Log (Average Precipitation (mm)) | -0.037 | 0.070 | -0.074** |
| p-value | (0.14) | (0.11) | (0.02) |
| Average O<sub>3</sub> Concentration (ppb) | -0.028*** | 0.009 | -0.041*** |
| p-value | (0.00) | (0.64) | (0.00) |
| Average PM2.5 Concentration (µg/m<sup>3</sup>) | 0.009 | 0.012 | 0.015 |
| p-value | (0.62) | (0.75) | (0.46) |
| Average UV Light Index | 0.161 | -0.243 | 0.185 |
| p-value | (0.22) | (0.52) | (0.24) |
| Set of Geographic and Time Controls: |  |  |  |
| Social Distancing Measures | Yes | Yes | Yes |
| County Fixed-Effect | Yes | Yes | Yes |
| Region-Level Day Fixed-Effect | Yes | Yes | Yes |
| County Time-trends | Yes | Yes | Yes |
| Number of Counties | 1,261 | 458 | 803 |
| Observations | 41,958 | 15,684 | 26,274 |
| R-squared | 0.114 | 0.151 | 0.111 |

*Notes:* Counties whose centroid is above the 39°50'N are considered northern. All standard errors are clustered at the county level. Numbers in parentheses are robust p-values. Signs \*\*\*, \*\*, and \* indicate 1%, 5%, and 10% statistical significance levels. The minimum and maximum temperatures are in Fahrenheit. Precipitation is in millimeters. O<sub>3</sub> is the maximum 8-hour concentration level measured in parts per billion, ppb. PM2.5 is the average daily concentration level measured in micrograms per cubic meters, µg/m<sup>3</sup>.

**Table A4.** Multiple regression results of estimating the association of weather and air quality indicators with the logarithm of county-level daily death rate per 18+ population in 1,281 counties and 43,480 county days - Average weather variables are calculated over a 3-day period: 19 through 21 days before death. *Interpretation of the reported coefficients:* Percentage (in decimals) change in daily death per 18+ county population for a one-unit change in the weather and air quality indicator.

| Variables | (1)<br>Model 1 | (2)<br>Model 2 | (3)<br>Model 3 | (4)<br>Model 4 | (5)<br>Model 4 | (6)<br>Model 4 | (7)<br>Model 4 | (8)<br>Model 4 |
| --- | --- | --- | --- | --- | --- | --- | --- | --- |
| Average Minimum Temperature (F) | 0.023*** | 0.014*** | 0.015** | 0.012* | 0.013** | 0.013* | 0.013* | 0.011 |
| p-value | (0.00) | (0.01) | (0.02) | (0.05) | (0.05) | (0.06) | (0.06) | (0.11) |
| Average Maximum Temperature (F) | -0.013*** | -0.007* | -0.000 | 0.003 | 0.002 | 0.002 | 0.002 | 0.000 |
| p-value | (0.00) | (0.09) | (0.95) | (0.56) | (0.70) | (0.70) | (0.74) | (0.95) |
| Log (Average Precipitation (mm)) |  |  |  |  | -0.009 | -0.009 | -0.007 | -0.005 |
| p-value |  |  |  |  | (0.53) | (0.53) | (0.65) | (0.73) |
| Average O<sub>3</sub> Concentration (ppb) |  |  |  |  |  | -0.000 | -0.000 | 0.001 |
| p-value |  |  |  |  |  | (0.96) | (0.93) | (0.89) |
| Average PM2.5 Concentration (µg/m<sup>3</sup>) |  |  |  |  |  |  | 0.008 | 0.007 |
| p-value |  |  |  |  |  |  | (0.37) | (0.43) |
| Average UV Light Index |  |  |  |  |  |  |  | 0.166* |
| p-value |  |  |  |  |  |  |  | (0.06) |
| Set of Geographic and Time Controls: |  |  |  |  |  |  |  |  |
| Social Distancing Measures | Yes | Yes | Yes | Yes | Yes | Yes | Yes | Yes |
| County Fixed-Effects | Yes | Yes | Yes | Yes | Yes | Yes | Yes | Yes |
| US-Level Day Fixed-Effects | Yes | Yes |  |  |  |  |  |  |
| Region-Level Day Fixed-Effects |  |  | Yes | Yes | Yes | Yes | Yes | Yes |
| County-Specific Time-Trends |  | Yes |  | Yes | Yes | Yes | Yes | Yes |
| R-squared | 0.023 | 0.096 | 0.046 | 0.113 | 0.113 | 0.113 | 0.113 | 0.113 |

*Notes:* All standard errors are clustered at the county level. Numbers in parentheses are robust p-values. Signs \*\*\*, \*\*, and \* indicate 1%, 5%, and 10% statistical significance levels. The minimum and maximum temperatures are in Fahrenheit. Precipitation is in millimeters. O<sub>3</sub> is the maximum 8-hour concentration level measured in parts per billion, ppb. PM2.5 is the average daily concentration level measured in micrograms per cubic meters, µg/m<sup>3</sup>.

**Table A5.** Multiple regression results of estimating the association of weather and air quality indicators with the logarithm of county-level daily deaths per 18+ population in northern and southern counties, using the preferred statistical model (Model 4) - Average weather variables are calculated over a 3-day period: 19 through 21 days before death. *Interpretation of the reported coefficients:* Percentage (in decimals) change in daily death per 18+ county population for a one-unit change in the weather and air quality indicator.

| Variables | Per 18+ County Population |  |  |
| --- | --- | --- | --- |
|  | Full Sample | Northern Sample | Southern Sample |
| Average Minimum Temperature (F) | 0.011 | 0.032** | -0.003 |
| p-value | (0.11) | (0.02) | (0.70) |
| Average Maximum Temperature (F) | 0.000 | -0.003 | 0.000 |
| p-value | (0.95) | (0.74) | (1.00) |
| Log (Average Precipitation (mm)) | -0.005 | 0.016 | -0.012 |
| p-value | (0.73) | (0.55) | (0.53) |
| Average O<sub>3</sub> Concentration (ppb) | 0.001 | 0.013 | -0.002 |
| p-value | (0.89) | (0.22) | (0.72) |
| Average PM2.5 Concentration (µg/m<sup>3</sup>) | 0.007 | 0.049** | 0.002 |
| p-value | (0.43) | (0.02) | (0.88) |
| Average UV Light Index | 0.166* | 0.037 | 0.162 |
| p-value | (0.06) | (0.88) | (0.13) |
| Set of Geographic and Time Controls: |  |  |  |
| Social Distancing Measures | Yes | Yes | Yes |
| County Fixed-Effect | Yes | Yes | Yes |
| Region-Level Day Fixed-Effect | Yes | Yes | Yes |
| County Time-trends | Yes | Yes | Yes |
| Number of Counties | 1,281 | 461 | 820 |
| Observations | 43,480 | 16,181 | 27,299 |
| R-squared | 0.113 | 0.150 | 0.109 |

*Notes:* Counties whose centroid is above the 39°50'N are considered northern. All standard errors are clustered at the county level. Numbers in parentheses are robust p-values. Signs \*\*\*, \*\*, and \* indicate 1%, 5%, and 10% statistical significance levels. The minimum and maximum temperatures are in Fahrenheit. Precipitation is in millimeters. O<sub>3</sub> is the maximum 8-hour concentration level measured in parts per billion, ppb. PM2.5 is the average daily concentration level measured in micrograms per cubic meters, µg/m<sup>3</sup>.

**Table A6.**
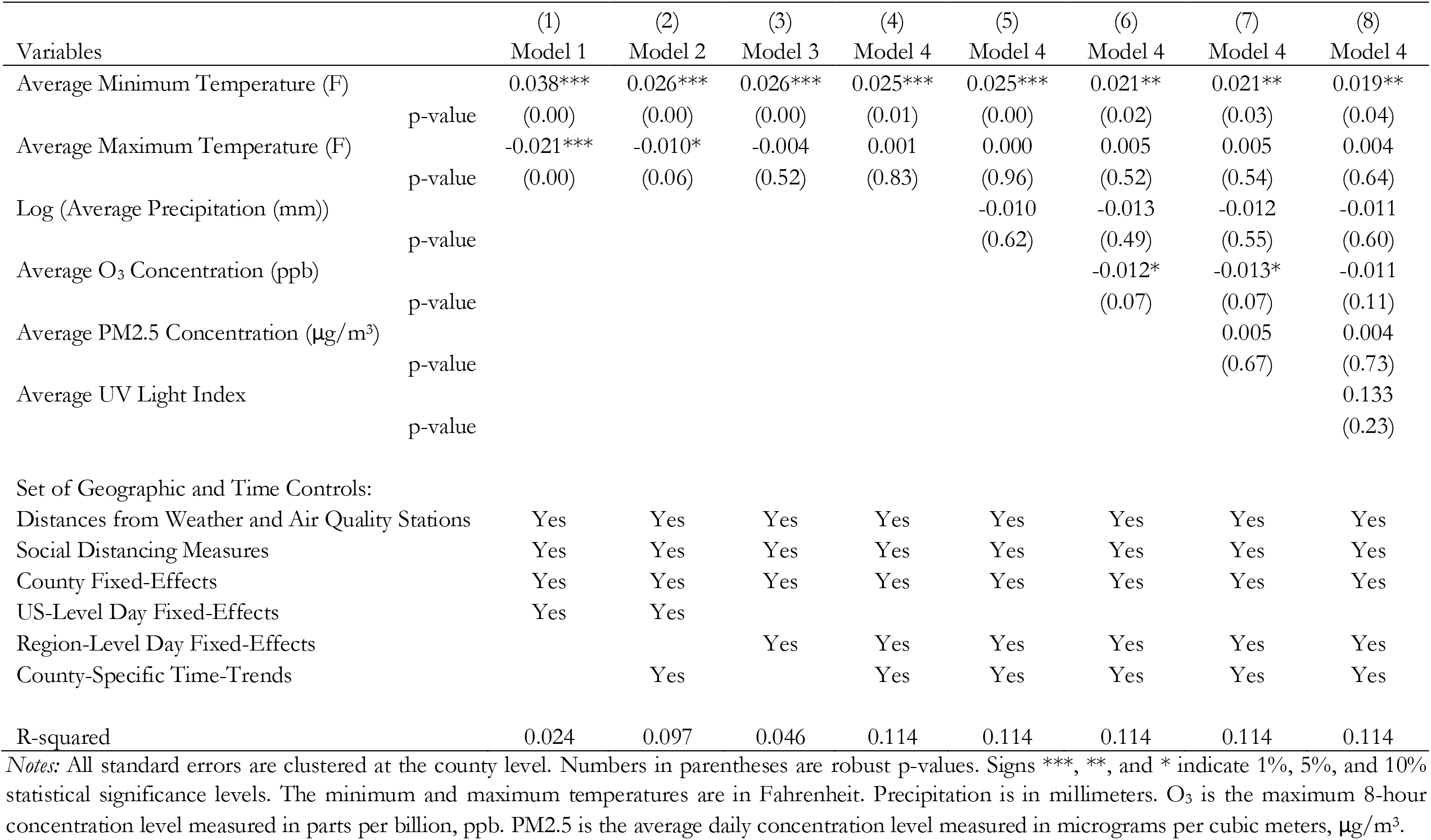
Multiple regression results of estimating the association of weather and air quality indicators with the logarithm of county-level daily death rate per 18+ population in 1,273 counties and 42,674 county days - Average weather variables are calculated over a 5-day period: 18 through 22 days before death, and distances from weather and air quality stations are added as covariates. *Interpretation of the reported coefficients:* Percentage (in decimals) change in daily death per 18+ county population for a one-unit change in the weather and air quality indicator.

| Variables | (1)<br>Model 1 | (2)<br>Model 2 | (3)<br>Model 3 | (4)<br>Model 4 | (5)<br>Model 4 | (6)<br>Model 4 | (7)<br>Model 4 | (8)<br>Model 4 |
| --- | --- | --- | --- | --- | --- | --- | --- | --- |
| Average Minimum Temperature (F) | 0.038*** | 0.026*** | 0.026*** | 0.025*** | 0.025*** | 0.021** | 0.021** | 0.019** |
| p-value | (0.00) | (0.00) | (0.00) | (0.01) | (0.00) | (0.02) | (0.03) | (0.04) |
| Average Maximum Temperature (F) | -0.021*** | -0.010* | -0.004 | 0.001 | 0.000 | 0.005 | 0.005 | 0.004 |
| p-value | (0.00) | (0.06) | (0.52) | (0.83) | (0.96) | (0.52) | (0.54) | (0.64) |
| Log (Average Precipitation (mm)) |  |  |  |  | -0.010 | -0.013 | -0.012 | -0.011 |
| p-value |  |  |  |  | (0.62) | (0.49) | (0.55) | (0.60) |
| Average O<sub>3</sub> Concentration (ppb) |  |  |  |  |  | -0.012* | -0.013* | -0.011 |
| p-value |  |  |  |  |  | (0.07) | (0.07) | (0.11) |
| Average PM2.5 Concentration (µg/m<sup>3</sup>) |  |  |  |  |  |  | 0.005 | 0.004 |
| p-value |  |  |  |  |  |  | (0.67) | (0.73) |
| Average UV Light Index |  |  |  |  |  |  |  | 0.133 |
| p-value |  |  |  |  |  |  |  | (0.23) |
| Set of Geographic and Time Controls: |  |  |  |  |  |  |  |  |
| Distances from Weather and Air Quality Stations | Yes | Yes | Yes | Yes | Yes | Yes | Yes | Yes |
| Social Distancing Measures | Yes | Yes | Yes | Yes | Yes | Yes | Yes | Yes |
| County Fixed-Effects | Yes | Yes | Yes | Yes | Yes | Yes | Yes | Yes |
| US-Level Day Fixed-Effects | Yes | Yes |  |  |  |  |  |  |
| Region-Level Day Fixed-Effects |  |  | Yes | Yes | Yes | Yes | Yes | Yes |
| County-Specific Time-Trends |  | Yes |  | Yes | Yes | Yes | Yes | Yes |
| R-squared | 0.024 | 0.097 | 0.046 | 0.114 | 0.114 | 0.114 | 0.114 | 0.114 |

*Notes:* All standard errors are clustered at the county level. Numbers in parentheses are robust p-values. Signs \*\*\*, \*\*, and \* indicate 1%, 5%, and 10% statistical significance levels. The minimum and maximum temperatures are in Fahrenheit. Precipitation is in millimeters. O<sub>3</sub> is the maximum 8-hour concentration level measured in parts per billion, ppb. PM2.5 is the average daily concentration level measured in micrograms per cubic meters, µg/m<sup>3</sup>.

**Table A7.** Multiple regression results of estimating the association of weather and air quality indicators with the logarithm of county-level daily deaths per 18+ population in northern and southern counties, using the preferred statistical model (Model 4) - Average weather variables are calculated over a 5-day period: 18 through 22 days before death, and distances from weather and air quality stations are added as covariates. *Interpretation of the reported coefficients:* Percentage (in decimals) change in daily death per 18+ county population for a one-unit change in the weather and air quality indicator.

| Variables |  | Per 18+ County Population |  |  |
| --- | --- | --- | --- | --- |
|  |  | Full Sample | Northern Sample | Southern Sample |
| Average Minimum Temperature (F) |  | 0.019** | 0.051*** | -0.003 |
|  | p-value | (0.04) | (0.01) | (0.81) |
| Average Maximum Temperature (F) |  | 0.004 | -0.001 | 0.003 |
|  | p-value | (0.64) | (0.93) | (0.74) |
| Log (Average Precipitation (mm)) |  | -0.011 | 0.042 | -0.028 |
|  | p-value | (0.60) | (0.21) | (0.27) |
| Average O<sub>3</sub> Concentration (ppb) |  | -0.011 | 0.007 | -0.018** |
|  | p-value | (0.11) | (0.63) | (0.03) |
| Average PM<sub>2.5</sub> Concentration (µg/m<sup>3</sup>) |  | 0.004 | 0.028 | 0.004 |
|  | p-value | (0.73) | (0.34) | (0.78) |
| Average UV Light Index |  | 0.133 | -0.117 | 0.100 |
|  | p-value | (0.23) | (0.71) | (0.45) |
| Set of Geographic and Time Controls: |  |  |  |  |
| Distances from Weather and Air Quality Stations |  | Yes | Yes | Yes |
| Social Distancing Measures |  | Yes | Yes | Yes |
| County Fixed-Effect |  | Yes | Yes | Yes |
| Region-Level Day Fixed-Effect |  | Yes | Yes | Yes |
| County Time-trends |  | Yes | Yes | Yes |
| Number of Counties |  | 1,261 | 458 | 803 |
| Observations |  | 42,674 | 15,931 | 26,743 |
| R-squared |  | 0.114 | 0.151 | 0.110 |

*Notes:* Counties whose centroid is above the 39°50'N are considered northern. All standard errors are clustered at the county level. Numbers in parentheses are robust p-values. Signs \*\*\*, \*\*, and \* indicate 1%, 5%, and 10% statistical significance levels. The minimum and maximum temperatures are in Fahrenheit. Precipitation is in millimeters. O<sub>3</sub> is the maximum 8-hour concentration level measured in parts per billion, ppb. PM<sub>2.5</sub> is the average daily concentration level measured in micrograms per cubic meters, µg/m<sup>3</sup>.

**Table A8.** Multiple regression results of estimating the association of weather and air quality indicators with the logarithm of county-level daily deaths per 18+ population in northern and southern counties, using state-level day fixed-effects *Interpretation of the reported coefficients:* Percentage (in decimals) change in daily death per 18+ county population for a one-unit change in the weather and air quality indicator.

| Variables | Per 18+ County Population |  |  |
| --- | --- | --- | --- |
|  | Full Sample | Northern Sample | Southern Sample |
| Average Minimum Temperature (F) | 0.011 | 0.045** | -0.012 |
| p-value | (0.34) | (0.03) | (0.41) |
| Average Maximum Temperature (F) | 0.009 | 0.005 | 0.012 |
| p-value | (0.35) | (0.76) | (0.28) |
| Log (Average Precipitation (mm)) | 0.029 | 0.085* | 0.004 |
| p-value | (0.25) | (0.06) | (0.89) |
| Average O<sub>3</sub> Concentration (ppb) | -0.003 | -0.009 | -0.005 |
| p-value | (0.71) | (0.63) | (0.66) |
| Average PM<sub>2.5</sub> Concentration (µg/m<sup>3</sup>) | 0.007 | 0.014 | 0.011 |
| p-value | (0.67) | (0.71) | (0.55) |
| Average UV Light Index | -0.098 | -0.432 | -0.074 |
| p-value | (0.60) | (0.40) | (0.72) |
| Set of Geographical and Time Controls: |  |  |  |
| Social Distancing Measures | Yes | Yes | Yes |
| County Fixed-Effect | Yes | Yes | Yes |
| State-Level Day Fixed-Effect | Yes | Yes | Yes |
| County Time-trends | Yes | Yes | Yes |
| Number of Counties | 1,273 | 460 | 813 |
| Observations | 42,674 | 15,931 | 26,743 |
| R-squared | 0.193 | 0.244 | 0.177 |

*Notes:* Counties whose centroid is above the 39°50'N are considered northern. All standard errors are clustered at the county level. Numbers in parentheses are robust p-values. Signs \*\*\*, \*\*, and \* indicate 1%, 5%, and 10% statistical significance levels. The minimum and maximum temperatures are in Fahrenheit. Precipitation is in millimeters. Ozone is the maximum 8-hour concentration level measured in parts per billion, ppb. PM<sub>2.5</sub> is the average daily concentration level measured in micrograms per cubic meters, µg/m<sup>3</sup>.

